# Prospective evaluation of ID NOW COVID-19 assay used as point-of-care test in an Emergency Department

**DOI:** 10.1101/2021.03.29.21253909

**Authors:** Jean-Claude Nguyen Van, Camille Gerlier, Benoît Pilmis, Assaf Mizrahi, Gauthier Péan de Ponfilly, Amir Khaterchi, Vincent Enouf, Olivier Ganansia, Alban Le Monnier

## Abstract

**Background:** Rapid testing for COVID-19 has been clearly identified as an essential component of the strategy to control the SARS-CoV-2 epidemic, worldwide. The ID NOW COVID-19 assay is a simple, user-friendly, rapid molecular biology test based on nicking and extension amplification reaction (NEAR).

**Objectives:** The aim of this study was to evaluate the ID NOW COVID-19 assay when used as a point-of-care test (POCT) in our Emergency Department (ED).

**Type of study:** This prospective study enrolled 395 consecutive patients; paired nasopharyngeal swabs were collected from each study participant. The first swab was tested with the ID NOW COVID-19 assay at the point-of-care by ED nurses. The second swab was diluted in viral transport medium (VTM) and sent to the clinical microbiology department for analysis by both the RT-PCR Simplexa test COVID-19 Direct assay as the study reference method, and the ID NOW COVID-19 assay performed in the laboratory.

**Results:** Nasopharyngeal swabs directly tested with the ID NOW COVID-19 assay yielded a sensitivity, specificity, PPV and NPV of 98.0%, 97.5%, 96.2% and 98.7%, respectively, in comparison with the RT-PCR study reference assay. When the ID NOW COVID-19 assay was performed in the laboratory using the VTM samples, the sensitivity decreased to 62.5% and the NPV to 79.7%. Three false negative test results were reported with the ID NOW COVID-19 assay when performed using undiluted swabs directly in the ED; these results were obtained from patients with elevated CT values (>30).

**Conclusion:** We demonstrated that the ID NOW COVID-19 assay, performed as a point of care test in the ED using dry swabs, provides a rapid and reliable alternative to laboratory-based RT-PCR methods

## Introduction

Coronavirus disease 2019 (COVID-19), caused by SARS-CoV-2, first appeared in China and then spread worldwide (1). The primary goal of the epidemic containment of COVID-19 is to reduce the infection transmission in the population by reducing the number of susceptible persons or by reducing the basic reproductive number (R0).

To date, the reference testing method is the real-time reverse transcriptase-PCR (RT-PCR) similar to that developed for the diagnosis of SARS-CoV in early 2000s (2). However, due to the rapid spreading of the SARS-CoV-2 pandemic and the limited capacities of molecular testing at the laboratory level, the concept of molecular testing in point-of-care setting such as in an Emergency Department (ED) appears to be useful to manage suspected infection cases. Indeed, the urgent need for increased testing for COVID-19 has been clearly identified as an essential component of the strategy to control the epidemic, worldwide.

The ID NOW COVID-19 assay is a simple, userfriendly, rapid molecular biology test and do not required specific equipment molecular biology, allowing point-of-care testing. This molecular isothermal assay is based on nicking enzyme amplification reaction (NEAR) technology that targets the *RdRP* gene (3) with a manufacturer’s claimed limit of detection (LOD) of 125 genome equivalents/mL. Results are provided in approximately 15 min, including upfront 3 minutes of sample elution buffer warm-up time. The assay run time on the ID NOW platform was within 2 to 5 min for a positive call in the early callout mode as opposed to less than 13 min on the full-callout mode for a negative result. To date, the reported analytical performance of the ID NOW COVID-19 test in the literature reviewed has been variable (4–6). This could be related to the conditions under which the samples were collected. The manufacturer’s instructions state that this test is valid only on direct swab and after discharged in VTM transport medium as well as limiting the number of days of symptoms to seven, otherwise the sensitivity would be drastically reduced (7, 8)

The objective of this work was first to evaluate the analytical performances of the ID NOW COVID-19 assay performed by ED nurses in point-of-care in comparison with the reference test RT-PCR Simplexa COVID-19 Direct assay (DiaSorin) currently in use in our laboratory. The second objective was to evaluate the effect of the transport medium VTM on the performance of the test.

## Materials and Methods

### Type of study

This prospective study was conducted between October 22 and December 22, 2020 in the ED of Groupe Hospitalier Paris Saint-Joseph, France. Consecutive adults presenting with clinical suspicion of COVID-19 assessed by the attending emergency physician and submitted to diagnostic tests for SARS-CoV-2 were eligible. The main clinical and laboratory data were collected in an electronic Case Report Form (e-CRF). All adult patients consenting to the participation in the study and to the use of biological samples were included. Patients were excluded when samples missed or inadequate.

### Clinical specimens

Paired nasopharyngeal swabs were collected with a flexible nasopharyngeal flocked swabs from patients having a clinical suspicion of COVID-19 by the attending nurse in the ED. Swabs were sent to the microbiology laboratory without prior assignment to either technique. The lack of identification of the order of collection and the size of the study ensures that the swabs were randomly distributed.

- One swab was directly used for the ID NOW COVID-19 assay in point-of-care test by ED trained nurses, previously trained and certified for use it. Training on the use of ID NOW was provided by qualified experts. Proficiency tests were conducted with ED nurses prior to study initiation. Testing was conducted according to instructions for use for ID NOW.
- The second swab collected, was then discharged in 3-ml viral transport medium (VTM) (Labo Moderne, Gennevilliers, France) and sent to the central microbiology laboratory for analysis.

Specimens were stored at 4°C up to a day, if all testing could not be completed on the same day. In case of discrepancies, samples were aliquoted and frozen at −80°C for confirmatory testing.

### Testing methods

The ID NOW COVID-19 assay (Abbott, Chicago, Il, USA) is an isothermal nucleic acid amplification-based. The assay was performed directly from the dry swab in the ED or by transferring 200 microliters of VTM to elution buffer in the sample base and then mixed for 10⍰seconds per instructions for use at the microbiology laboratory.

The Simplexa COVID-19 direct assay (Diasorin, Saluggia, Italy) was chosen as RT-PCR reference test and was performed with the DiaSorin LIAISON^®^ MDX according to the manufacturer’s instructions for use. A 50-μl volume of Simplexa COVID-19 Direct kit reaction mix (MOL415 0) was added to the “R” well of the 8-well direct amplification disc (DAD) followed by addition of 50⍰μl of non extracted nasopharyngeal swab (NPS) sample to the “SAMPLE” well. Fluorescent probes are used together with corresponding forward and reverse primers to amplify two different regions of the SARS-CoV-2 genome: *ORF1ab* and *S* gene. Data collection and analysis were performed with LIAISON^®^ MDX Studio software. *C*_*T*_ values were collected from MDX software.

### Confirmation of discrepant results

In case of discrepancy, a control was performed using an aliquot from VTM previously stored at –80 °C for quantitative RT-PCR following the protocol established by the National Reference Center for Respiratory Viruses (NRC) at Institut Pasteur, Paris, France (9, 10)

### Statistical analysis

The percentages were calculated based on documented data (missing data were excluded from the percentages). Inter-group comparisons were made using the Mann-Whitney test for quantitative variables and the Fisher exact test for qualitative variables. Statistical analysis was done with StatView software (version 5.0). All tests were two-tailed and p values less than 0.05 (calculated by *c* test, Student’s t test, or Mann-Whitney test) were considered significant.

### Ethical statement

This study followed the Standards for Reporting Diagnostic Accuracy study (STARD) guidelines and was favourable approved by the local clinical ethic committee board called “Comité de protection des Personnes Nord Ouest” N° IRB 2020-A02758-31. Informed oral consent for participation was obtained from each participant, in accordance with French law.

## Results

A total of 406 patients were enrolled in the study, mean age was 71 years and M/F sex ratio = 1.12. Among them, 11 were secondary excluded due to a lack of adequate specimen samples (Figure 1: Study Flow Chart). A total of 395 patients were eligible for study inclusion. Among these, 154 patients (39%) were diagnosed with SARS-CoV-2 infection (Figure 1).

**Figure 1.**
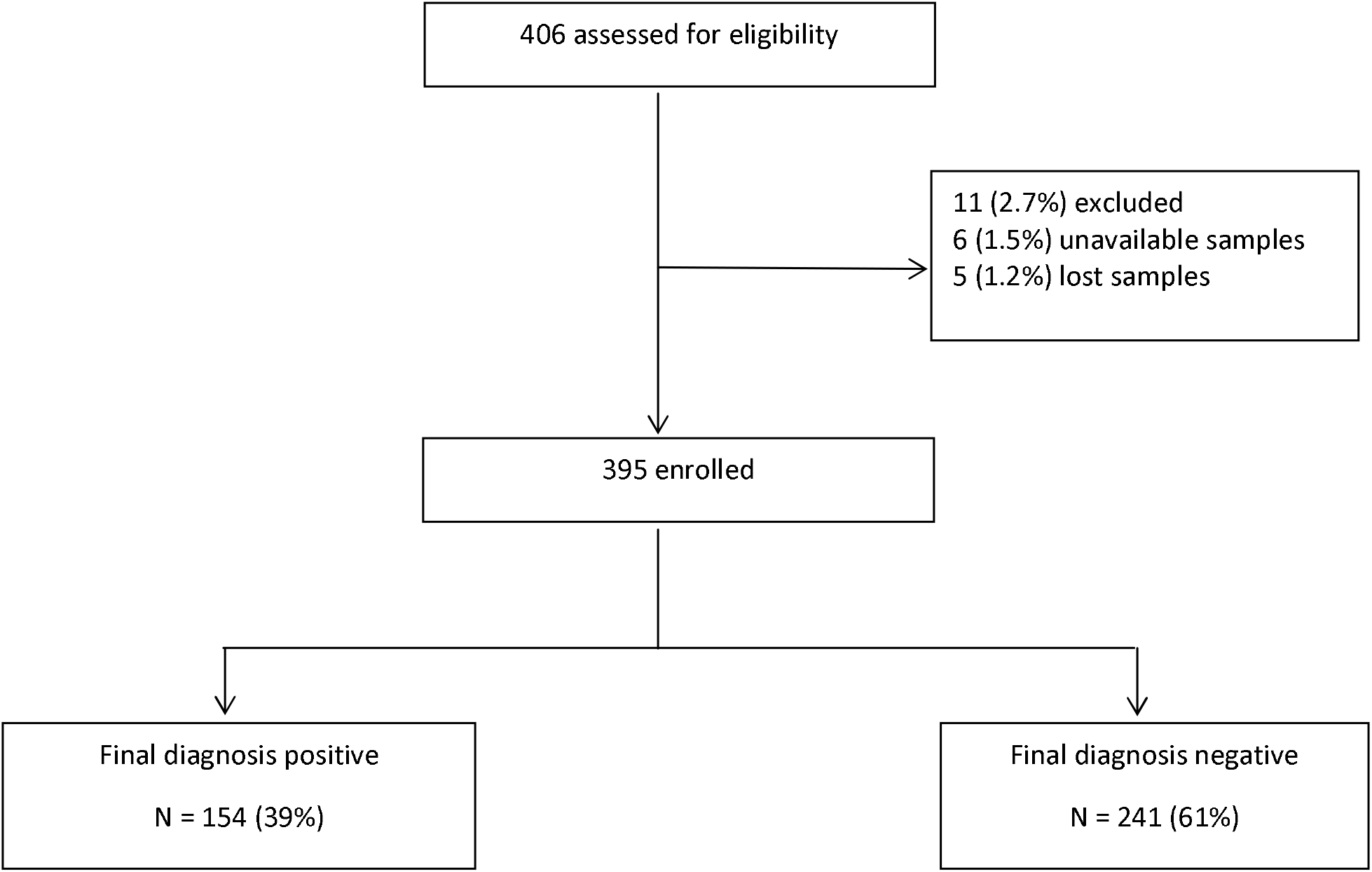
Study flow chart. The arrow indicate the study flow, up to the final classification of 154 patients with the infection and without the infection

### Performances of the ID NOW COVID-19 assay compared to reference RT-PCR assay

POCT in the ED with nasopharyngeal swabs directly tested using the ID NOW COVID-19 assay showed sensitivity, specificity, positive predictive value (PPV), and negative predictive value (NPV) of 98.0%, 97.5%, 96.2% and 98.7%, respectively. When the ID NOW COVID-19 assay was performed in the laboratory from the swab previously discharged in VTM, the sensitivity dropped to 62.5% and the NPV to 79.7% (Table 1). Three false negative test results were reported with the ID NOW COVID-19 assay when performed using undiluted swabs directly in the ED; these results were obtained from patients with elevated C_*T*_ values (>30).

**Table 1.**
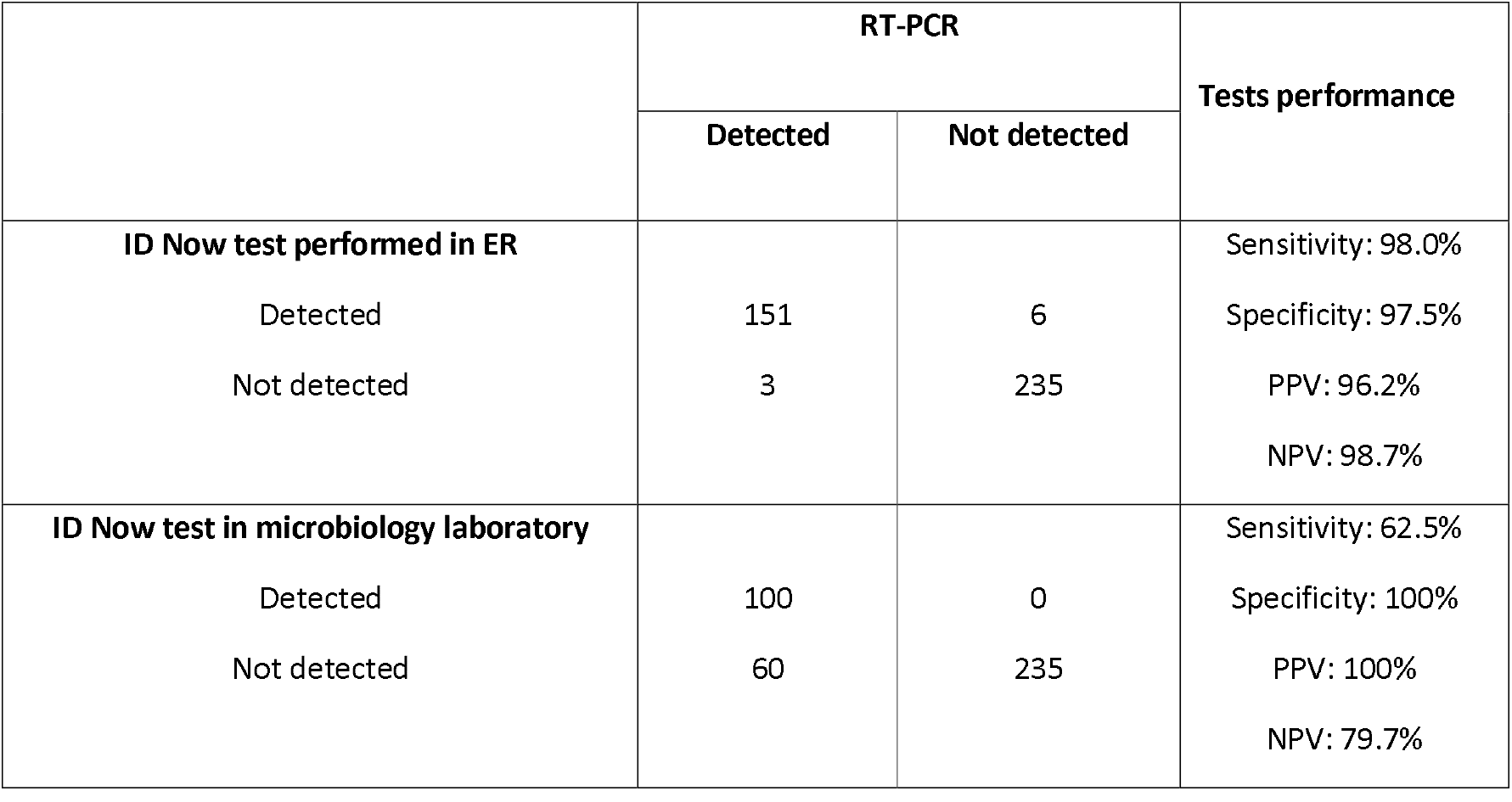
Analytical performances of the COVID-19 ID NOW test assay performed in emergency department and ID Now test in microbiology laboratory compared to reference RT-PCR assay

### Comparison of C_T_ values among positive samples detected by direct ID NOW COVID-19 assay and samples performed on VTM by ID NOW COVID-19 assay

The median cycle threshold (C_*T*_) for positive sample by direct ID NOW COVID-19 assay and ID NOW COVID-19 assay performed on VTM were 17.9 (interquartile range [IQR] [15.5 – 21.3]) whereas for positive sample only by direct ID NOW COVID-19 assay median (C_*T*_) were 30.7 [28.2 – 32.4]; p < 0.001 (Figure 2). Overall agreement between direct ID NOW and ID NOW performed on VTM were 66.2%.

**Figure 2.**
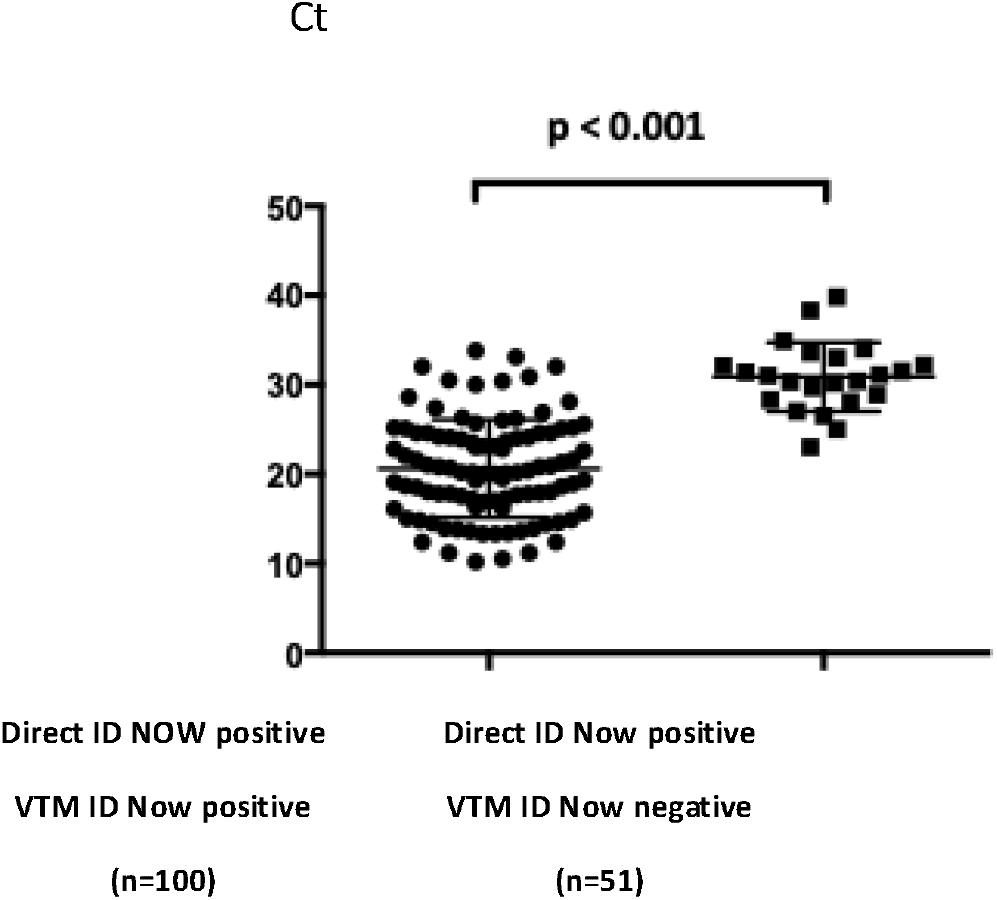
Comparison of C_*T*_ values among positive samples detected by direct ID NOW™ COVID-19 assay and ID NOW™ COVID-19 assay performed on VTM. C_*T*_ value differences were compared by using the Kruskal-Wallis test. VTM: viral transport medium

### Analysis of discordant results

We observed nine discrepancies between the results with the ID NOW COVID-19 assay performed directly in the ED and the RT-PCR method by Simplexa at the central laboratory (Table 2). An aliquot from VTM were sent to the National Reference Center for respiratory viruses at Institut Pasteur, Paris. Finally, we reported three samples (ID NOW COVID-19 assay negative, positive by RT-PCR) and six samples (ID NOW COVID-19 assay positive, control negative by RT-PCR).

With the three false negative samples, we made two observations: either a lack of detection of a low viral load SARS-CoV-2 infection (n=2) or a previous history of SARS-CoV-2 infection (n=1). Two patients presented with symptoms suggestive of SARS-CoV-2 infection. One had pneumonia with no respiratory severity criteria associated with a biological inflammatory syndrome and lymphopenia. The other patient had acute viral gastroenteritis with neither haemodynamic severity criteria, biological inflammatory syndrome, nor lymphopenia. The Simplexa MDX RT-PCR assay was performed within 24 hours of collection and positive result with a high C*t* in both cases (>30), indicating a low viral load.

The third patient had a history of COVID-19 six weeks earlier in a pauci-symptomatic form. She was being managed in the ED for a bowel obstruction and had no symptoms of COVID-19. However, indirect biological signs were present (inflammatory syndrome and lymphopenia). Simplexa MDX RT-PCR control and National Reference Center for Respiratory Viruses (NRC) control confirmed the persistence of low viral load with high C_*T*_ (>30) related to the previous infection. Heterogeneity of viral load between the two swabs may explain these discrepancies. Among the six false-positive swabs, we were also able to make two observations: known previous SARS-CoV-2 infections (n=3) and early forms of SARS-CoV-2 infections (n=3). Among the three patients with a history of COVID-19, two had been hospitalized for reasons unrelated to and distant from COVID-19 (8 weeks and 8 months after). The third had been hospitalized three weeks after the diagnosis of COVID-19 for an episode of dyspnea. Thoracic angioscanner found minimal (10-25%) sequential lung damage with no pulmonary embolism. Simplexa, and NRC Pasteur RT-PCR controls showed negative results which may be explained by heterogeneity of loading between the first and second nasal swab. Among the three patients with no history of COVID-19, timing context and associated symptoms were consistent with frequent modes of COVID-19 disclosure in the elderly: fall, anorexia, weakness and delirium. We hypothesize that these cases were early SARS-CoV-2 infections not detected by the RT-PCR controls because of a viral load defect on the second swab.

## Discussion

Rapid and accurate detection of SARS-CoV-2 is essential to ensure early and appropriate patient management, outbreak containment, and to better follow the global epidemiology of the virus. Laboratory testing is mainly based on amplification and detection of viral gene sequences in upper respiratory tract specimens. The results of these analyses are often lengthy (1-4 hours) and difficult to manage with a large flow of patients in the ED and lead to a risk of overcrowding in these units. As reported by Alter et *al*. (11) and our team (12), point-of-care test (POCT) instruments in ED produce lab results with rapid turnaround times.

This study demonstrated that the detection of SARS-CoV-2 using rapid isothermal nucleid acid amplification assay performed by nurses directly at the emergency room, 24 hours-a-day, 7 days-a-week was both feasible and reliable. The sensitivity and specificity of the ID NOW system for the detection of SARS-CoV-2 was higher than 98% even when performed by a trained professional (in this case a nurse) without direct laboratory supervision. Less than 3.8%of tests failed were observed during this study. As reported by Kanwar et al.(13), the invalid rate is an important consideration when selecting an assay for clinical use. Our experience with the ID NOW platform for the diagnosis of seasonal influenza showed satisfactory results (14) which allowed implementation POCT in our ED (12). This experience has enabled the rapid implementation of a POCT’s strategy in ED for the diagnosis of COVID-19.

We have to keep in mind that identification of the virus is only a part of clinical diagnosis and management. Although our results are important, further studies are necessary to assess the clinical impact of ID NOW COVID-19 assay at triage in order to evaluate its effect on decision-making and the prescription of complementary examinations. POCT is only useful if it has an impact on treatment or healthcare organization. Some studies conducted during influenza outbreaks have shown a positive impact on medical costs (15, 16), rapid antiviral treatment (17) and reduced length of stay (12), especially for patients with a negative result, highlighting the importance of tests that have a high negative predictive value. However, we demonstrated the significant decrease in sensitivity of the ID NOW COVID-19 assay when performed using diluted swabs in VTM. We strongly recommend to perform immediately the test on fresh swab only. Dilution of the nasopharyngeal swab specimen in VTM below the lower limit of detection for the assay increases the risk of false negative results. Thus, as reported by Basu *et al* (8) ID NOW COVID-19 assay is not optimized for testing nasopharyngeal swabs in VTM but the best performance would be obtained from dry nasal swabs and tested directly at the point-of-care. However, once the swab is discharged into the sample receiver, if the result is invalid, it is no longer usable for central laboratory test. In the case of invalid or negative result with clinical signs suggestive of SARS-CoV-2 infection, the physician should perform an RT-PCR confirmatory test. Our study has several limitations. First, the ID NOW COVID-19 assay itself has only one target, the *RdRP* gene. To date, the published isothermal amplification technology tests did not evaluate SARS-CoV-2 using two target genes (18). Second, this work was conducted during an epidemic peak and so the results should be viewed in the context of the conditions during such a period. Positive predictive values decrease when tests are performed with a lower prevalence of the disease, thus the results cannot be extrapolated to a lower prevalence period. Third, this study was conducted on the relatively small number of samples in a single care center setting. Therefore the results may be specific to the population of this center and may not be applicable to another patient group such as children for example. Use of rapid diagnostic test at triage in other care settings would depend on the organization of each ED and, importantly, on the availability of compliant and trained staff.

## Conclusion

With large numbers of patients being admitted to hospital, putting tremendous pressure on health care systems, there is an urgent demand for a user friendly, rapid, simple and sensitive POCT assay. Such tests could be used at hospitals to facilitate faster detection of SARS-CoV-2, which can reduce or avoid further spread and ensure appropriate patient management. This is the first prospective evaluation of ID NOW COVID-19 assay in “real life” in ED and we showed ID NOW COVID-19 assay used as POCT in an ED setting provides a rapid and reliable alternative to laboratory-based RT-PCR methods.

## Data Availability

None

## Acknowledgements

The authors would like to thank the Emergency Department team and especially Anne Boureau, Aurélie Lebigre, Trinidad Rodriguez, Bernadette Gauchet-Jegier, Mélanie Luce for the coordination within the ward and all ED nurses to have done all samples.

Special thanks to Tanguy Robardet and Salim Kahlat for technical support and the contribution of Olivier Billuart as data manager.

We are also grateful to the members of Clinical Research: Center: Estelle Plan, Julien Fournier, Dr Nesrine Ben Nasr, Dr Hélène Beaussier and Prof. Gilles Chatellier.

We thank Dr Gregory Destras at the Lyon National Reference Center for Respiratory Viruses (France) for helpful discussion.

We have no conflicts of interest to declare.

